## Supplementary figures for "Causal associations between plasma proteins and prostate cancer: a Proteome-Wide Mendelian Randomization"

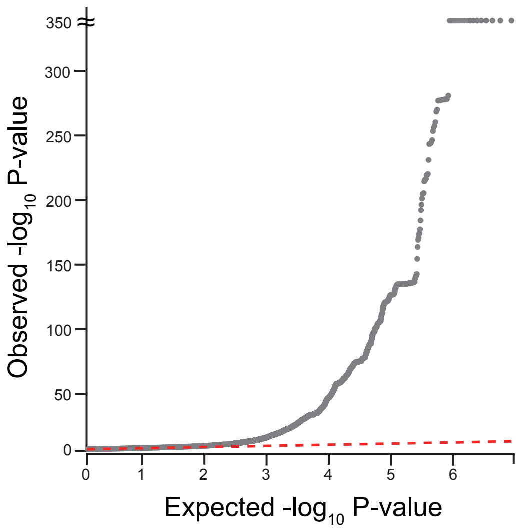


Supplementary Figure 1. Quantile-quantile plot of the PCa GWAS. The quantile-quantile plot illustrates the discrepancy between the actual P values of GWAS SNPs and the expected P values from a theoretical $\chi^2$ distribution.

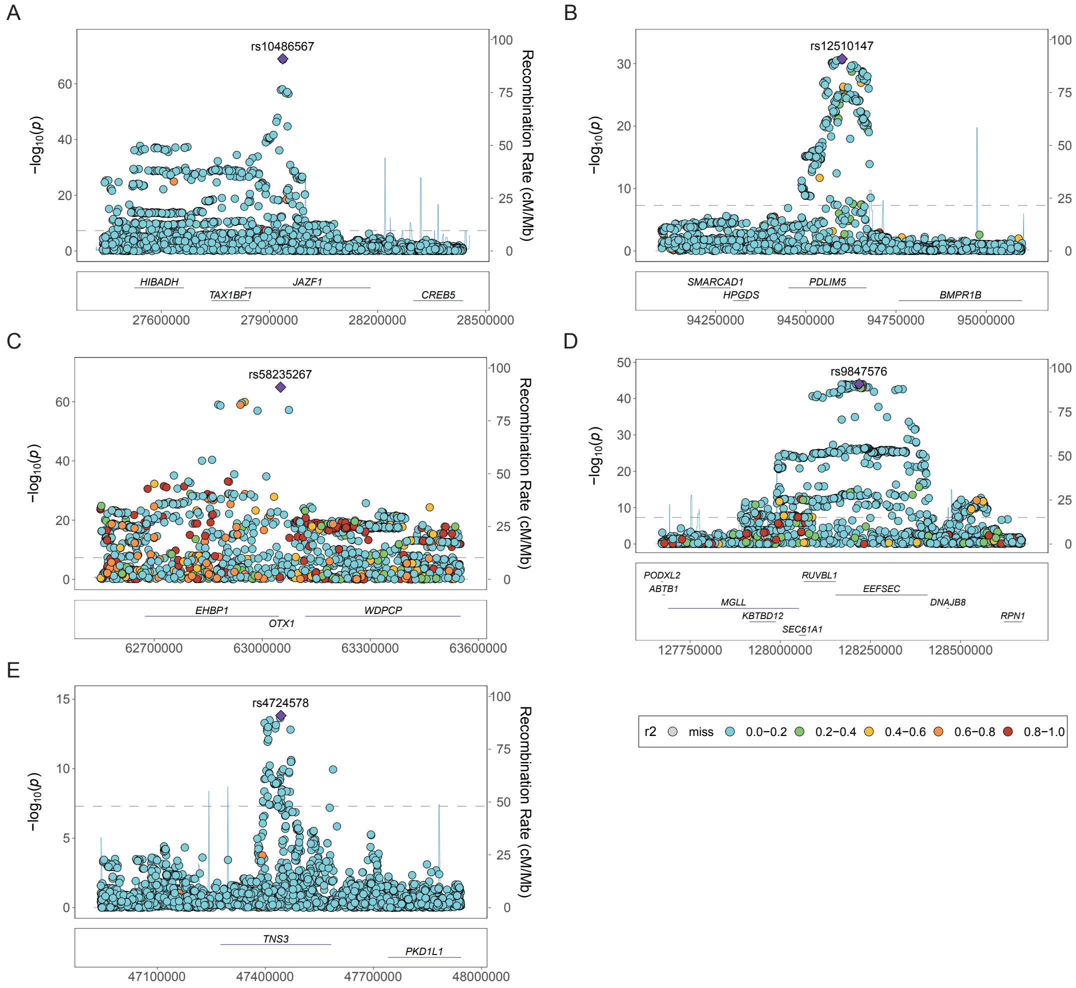


Supplementary Figure 2. LocusZoom plots of GWAS top SNPs. Genetic loci harboring top SNPs at the (A) JAZF1, (B) PDILM5, (C) WDPCP, (D) EEFSEC, and (E) TNS3 loci are displayed. LD value with the top SNP is represented by colors spanning from dark blue (low) to red (high). The dashed gray line represents the genome-wide significance threshold of $5\times10^{-8}$. Neighboring genes are shown at the bottom of the figure.

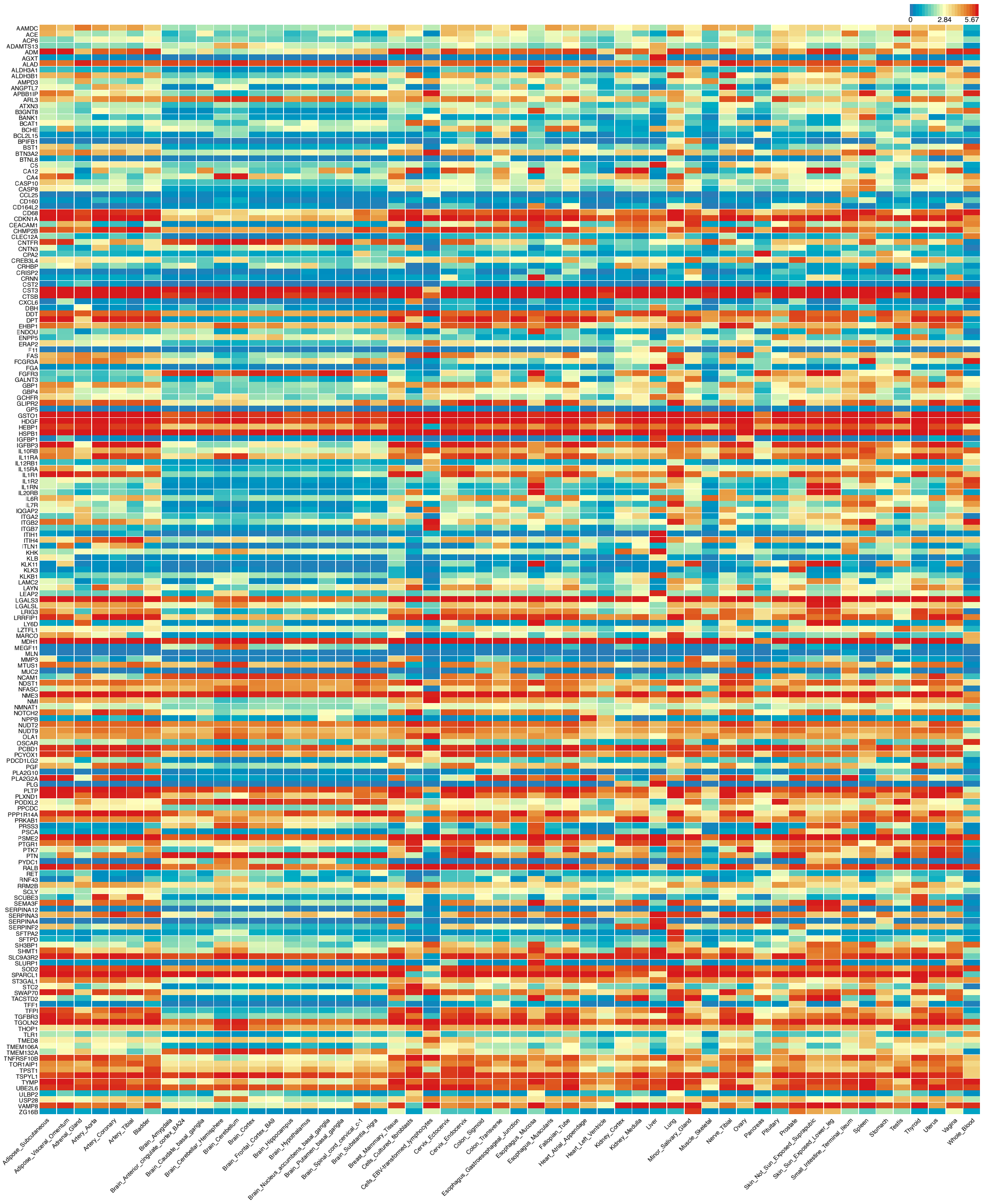

Supplementary Figure 3. Gene expression heat map of the 193 unique PW-MR significant proteins in deCODE and UKB-PPP. Log2-transformed average expression in 54 GTEx v8 tissues.
